# Sublingual Buprenorphine Is Associated with Increased Caries Experience and Altered Oral Microbial and Salivary Risk Factors

**DOI:** 10.64898/2026.11.13.26368056

**Authors:** Kyle V. Kasparian, Callahan Katrak, John Abernethy, Sydney Reed, Kyra Martin, Renita Jenkins, Yanan Zang, Shu-Hao Hsu, Dayane Oliveira, Bruna Garcia, Abhisheak Sharma, Christopher R. McCurdy, Jacqueline Abranches

## Abstract

**Background:** Sublingual buprenorphine (SLB) is commonly prescribed to treat opioid use disorder. Pharmacoepidemiologic studies suggest an association between SLB and severe dental caries, yet clinical investigations are lacking.

**Objective:** To assess caries experience and caries-related risk factors among patients taking SLB.

**Methods:** This pilot cross-sectional study enrolled 11 subjects taking SLB films, 10 taking SLB tablets, and 10 age- and sex-matched controls. Demographic, medical, dental, and behavioral data were collected. Caries examinations were performed. Salivary pH, buffering capacity, and flow rate were measured. Saliva and plaque levels of Mutans streptococci and *Candida* spp. were determined. Oral pH dynamics and salivary concentrations of buprenorphine, its metabolites, and naloxone were monitored.

**Results:** Caries experience was greater in SLB subjects than controls and higher among tablet versus film subjects (p<0.05). SLB subjects exhibited lower salivary pH and buffering capacity (p<0.05), and elevated salivary levels of Mutans streptococci (p<0.05) and *Candida* spp. (p≤0.001). SLB administration was associated with rapid oral acidification and prolonged recovery. Salivary buprenorphine exposure was elevated and correlated with baseline pH (p<0.01), suggesting ion trapping contributes to oral retention.

**Conclusion:** Patients receiving SLB exhibit increased caries experience likely resulting from localized acidification favoring caries-associated microorganisms and prolonged oral exposure to buprenorphine.

## Introduction

Opioid use disorder (OUD) is a chronic, debilitating condition affecting millions of Americans, and carries an estimated economic burden exceeding $700 billion^1^. Administered as a film or tablet, sublingual buprenorphine (SLB) is the most commonly prescribed OUD medication^2^ as it effectively prevents withdrawal and relapse^3^. While buprenorphine has improved millions of lives, only ∼25% of the OUD community utilize pharmacotherapy^4^, making patient acceptance and adherence to effective drug products critical.

In 2022, the FDA released a warning that oral transmucosal drug products containing buprenorphine are associated with increased dental decay (i.e., caries) and tooth loss^5^. The warning has garnered many patients’ attention, which could diminish acceptance and adherence to SLB. The warning is supported by pharmacoepidemiologic evidence that found a strong association even when controlling for indication (i.e., OUD patients not taking SLB) or when controlling for systemic buprenorphine exposure (i.e., patients taking transdermal buprenorphine indicated for chronic pain)^6–10^. This suggests a unique unknown mechanism regarding SLB and dental caries. Recent preclinical evidence suggests that salivary glands act as a depot for buprenorphine, with sublingual administration driving chronically elevated levels in the oral cavity that could increase biofilm accumulation and risk of caries^11,12^.

Dental caries is a multifactorial, polymicrobial biofilm-based disease that results in dissolution of the tooth structure and eventual cavitation by microbially produced organic acids. Frequent and prolonged acidification of dental plaque favors highly aciduric microorganisms, inducing dysbiosis which drives caries initiation and progression^13^. *Streptococcus mutans* is a key cariogenic pathogen, as it forms robust biofilms that adhere to the tooth and rapidly generates organic acids from fermentable carbohydrates^14^. Additionally, *Candida* species, particularly *Candida albicans*, are often co-isolated with *S. mutans* from aggressive carious lesions^15,16^. Importantly, patients with hyposalivation, decreased resting salivary pH, and/or decreased salivary buffering capacity have an impaired ability to neutralize acids, and therefore have increased caries risk^17^.

Here, we conducted the first cross-sectional clinical pilot study to assess caries experience, microbial burden, salivary factors, and oral pH dynamics in SLB subjects by formulation (i.e., films vs. tablets). We also quantified buprenorphine, its major metabolites, and naloxone in saliva post-dose to characterize salivary pharmacokinetics and evaluate how subject-specific salivary properties may influence retention in the oral cavity. Ultimately the results of this study reveal novel insights into the contributory mechanisms explaining the association of SLB use with dental caries.

## Methods

### Participant Recruitment and Enrollment

This cross-sectional clinical study was approved by the University of Florida Institutional Review Board (IRB202401848). Given the absence of prior clinical data to inform a reliable effect-size estimate, this study was designed as an exploratory pilot. Similar to other studies^18^, the pragmatic enrollment of 30 participants was selected, which could inform the design and sample-size calculation of future adequately powered studies. Subjects were primarily recruited from surrounding addiction medicine clinics. We enrolled 21 subjects maintained on SLB ≥1 year. Eleven subjects were prescribed films while ten were prescribed tablets. We additionally recruited and enrolled 10 age- and sex-matched subjects with no prior diagnosis of OUD and no history of SLB use as controls. Eligibility was confirmed per Table S1. Participants abstained from oral hygiene overnight and consumed no food/beverages except water for two hours before the visit. All participants completed and signed informed consent.

### Survey Data Collection

Subjects completed a questionnaire assessing demographics, general medical history, dental history, oral hygiene practices, and dietary consumption. This questionnaire was derived from the Caries Management by Risk Assessment (CAMBRA), a validated dental caries risk assessment tool^19^. Dietary sugar intake was assessed using a predefined list, and frequency was categorized by highest reported intake.

### Clinical Examination and Sample Collection

Clinical examinations were conducted by a single dentist (CK). Caries detection and diagnosis followed the modified International Caries Detection and Assessment System II (ICDAS-II) including lesion activity assessment^20^. All clinical examinations were performed by a single dentist (CK) who completed several calibration exercises, ensuring standardized application of ICDAS-II diagnostic criteria throughout the study.

The number of decayed (to dentin), missing, and filled teeth (DMFT) and surfaces (DMFS) was calculated for each subject. Third molars were excluded, yielding a denominator of 28 teeth. Root surfaces were included, so six surfaces per tooth were assessed (168 surfaces total). Missing teeth due to caries were identified through patient self-report of extraction due to dental caries or were assumed to be caries-related unless a non-caries reason (e.g., orthodontic treatment, congenitally missing, or periodontal disease) was known. Supragingival dental plaque level was qualitatively assessed by a single dentist (CK), and categorized into three levels: minimal, moderate, or high if < 25%, 25-50%, or > 50% of tooth surfaces had dental plaque present, respectively.

Unstimulated saliva was collected from subjects into sterile collection tubes for at least ten minutes, and salivary flow rate (USFR) was calculated. Supragingival dental plaque was collected using a sterile dental instrument from a non-carious enamel surface and carious tooth surface when present and placed into a tube containing 50 μL sterile deionized water. Plaque was preferentially collected from mandibular teeth. Plaque collected from carious surfaces was preferentially collected from dentin lesions.

The pH of each sample type was recorded using a calibrated micro pH probe (LabSen 241-3SP, Apera Instruments). 25 µL of plaque samples were immediately dispersed in 475 μL of sterile phosphate-buffered saline (PBS) and stored on ice until processing. Salivary buffering capacity was determined using Saliva-Check BUFFER kit (GC America).

### Assessing Microbiological Burden and Detection

Samples were processed as previously described in our laboratory^15,16^. Briefly, serial dilutions of dental plaque from both a non-carious enamel surface and a carious tooth surface and saliva samples were prepared, and 0.10 mL was plated on Tryptic Soy Agar (TSA), Sabouraud agar supplemented with 0.4 mg/ml chloramphenicol (SAB), and Difco Mitis Salivarius agar supplemented with 0.388 mg/ml bacitracin (MSB). MSB plates were incubated for 48 hours at 37 °C with 5% CO_2_ while SAB plates were incubated for 48 hours in aerobic conditions at 37 °C. TSA plates were first incubated for 24 hours at 37 °C with 5% CO_2_, then moved to aerobic conditions at 37 °C for another 24 hours to allow for growth of aerobic microorganisms. TSA is a nonselective media, thereby representing levels of total cultivable microorganisms. To quantify relative *Candida* spp. burden (Percent *Candida* spp./total microorganisms), colony forming units on SAB were counted and divided by 0.10 mL (volume plated) to yield CFUs/mL. This value was normalized to the CFUs/mL calculated from TSA plates. Similarly, to quantify relative Mutans streptococci (Percent MS/total microorganisms), CFUs/mL on MSB were normalized to CFUs/mL on TSA.

To detect *S. mutans*, polymerase chain reactions (PCR) of eight S. *mutans*-like colonies from MSB plates were performed as previously described^16^. Briefly, cells were lysed and PCR reactions were prepared using *S. mutans*-specific primers^21^ and GoTaq^®^ G2 Green PCR master mix (Promega, Madison, WI, USA). *S. mutans* laboratory strain, UA159, was used as a positive control for all PCR reactions. All reactions were run on agarose gel electrophoresis using Invitrogen SYBR Safe DNA gel stain (Thermo Fischer Scientific, Waltham, MA, USA).

To detect *C. albican*s, eight *Candida* spp. Colonies from SAB plates were streaked on CHROMagar™ Candida (Santa Maria, CA, USA), a chromogenic agar plate, for identification of select *Candida* species (*C. albicans, C. tropicalis, C. glabrata and C. krusei*). While green colonies are intended to selectively identify *C. albicans*, they may represent either *C. albicans* or *C. dubliniensis*^22^. Given its markedly higher prevalence in the oral cavity^23^, we classified all green colonies as *C. albicans*.

### Oral pH Kinetics and Disintegration Time

For participants taking SLB, saliva and plaque samples were collected 5, 10, 15, and 30 minutes after self-administration. Samples were collected as described in Supplemental Materials, and pH recorded. Saliva samples were then stored on ice for downstream bioanalysis. Caries-associated pH parameters including the minimum pH (pH_min_), area below critical pH (Area < pH_c_), and time below critical pH (Time < pH_c_) were calculated using GatorpH software^24^. Disintegration time was defined as visible disappearance of film or tablet by a study team member.

### Quantification of buprenorphine, norbuprenorphine, buprenorphine-glucuronide, norbuprenorphine-glucuronide, and naloxone in saliva

A bioanalytical method was developed using ultra-performance liquid chromatography-tandem mass spectrometry (UPLC-MS/MS) as detailed in the Supplemental Materials.

### Data analysis

Differences in the distributions of categorical demographic, clinical, and behavioral variables were assessed using Pearson chi-square tests. When overall chi-square tests were significant, Agresti–Finlay adjusted standardized residuals were examined to identify which specific group–category combinations contributed to the observed differences. Continuous numerical data was compared between groups using multiple Mann-Whitney tests. The p-values reported were adjusted using the false discovery rate (FDR) approach and a two-stage step-up method of Benjamini, Krieger, and Yekutieli.

Multiple chi-square tests with pairwise comparisons, random forest analysis, and multiple linear regression analyses were performed using RStudio version 2026.09.0+174. All other analyses were performed using GraphPad Prism 10 (GraphPad Software, San Diego, CA, USA).

## Results

### Analysis of demographics and caries risk modifiers

Thirty-one subjects were enrolled. One subject in the film group was excluded only from caries assessment and oral risk factor analysis due to the participant switching from tablet to film recently, while a different subject in the film group was excluded from our salivary pharmacokinetics analysis due to prematurely swallowing the film during administration. Therefore, a total of ten subjects per group were analyzed.

Our study population was age- and sex-matched, with no significant differences in demographics. Participants taking films or tablets were more likely to use tobacco than the controls (p<0.05), and subjects taking tablets were also more likely to have previously used cannabis (p<0.01) or other non-opioid illicit substances (p<0.01) versus controls (Table 1). The only significant difference between film and tablet groups was prior cannabis use (p<0.05). Tablet subjects had a higher qualitative plaque level versus controls (p<0.05) (Table 1). There were no significant differences found in frequency of brushing, flossing, or intake of high-sugar dietary items. However, this does not disprove the potential for differences that were not statistically detectable due to small sample size. Participants taking SLB were asked additional questions summarized in Table S2.

**Table 1:** Overview of demographics, prior substance use, diet, and oral hygiene practices across groups. . A total of 31 subjects were enrolled. One film subject was excluded from analysis due to recently switching from film to tablet, and therefore a total of 10 subjects per group were analyzed. Multiple Mann Whitney tests performed to compare age distributions. For categorical variables, Pearson chi-square tests were performed. When the overall chi-square test was significant, Agresti-Finlay adjusted standardized residuals were calculated to identify significant cell-level differences. Asterisks (*) represent statistically significant comparisons relative to the control group: *: p-value < 0.05; **: p-value < 0.01. Hashes (^#^) represent statistically significant comparisons between the film and tablet groups: ^#^: p-value < 0.05.

| Characteristics |  | No. |  |  |
|---|---|---|---|---|
|  |  | Controls | Film Group | Tablet Group |
| Age, range (median), years |  | 23 – 61 (41) | 28 – 78 (42) | 27 – 66 (43.5) |
| Sex | Male | 3 | 2 | 5 |
|  | Female | 7 | 8 | 5 |
| Race/Ethnicity | White | 7 | 9 | 8 |
|  | Asian | 2 | 0 | 0 |
|  | Black | 1 | 1 | 1 |
|  | American Indian, Hispanic | 0 | 0 | 1 |
| Education Level | Grade School | 0 | 1 | 1 |
|  | High School | 2 | 4 | 5 |
|  | Trade School | 0 | 2 | 0 |
|  | College | 8 | 3 | 4 |
| Tobacco Use |  | 0 | 5* | 5* |
| Cannabis Use | Current | 0 | 3 | 2 |
|  | Past Use | 1 | 1 | 7**<sup>, #</sup> |
| Illicit Substance Use | Current | 0 | 2 | 0 |
|  | Past Use | 0 | 1 | 7** |
| Plaque Score | Minimal | 7 | 3 | 1* |
|  | Moderate | 3 | 5 | 5 |
|  | Heavy | 0 | 2 | 4* |
| Brushing Frequency | No daily brushing | 0 | 1 | 1 |
|  | 1x/day | 4 | 3 | 4 |
|  | 2x/day | 6 | 6 | 5 |
| Flossing Frequency | No daily flossing | 3 | 6 | 8 |
|  | ≥ 1x/day | 7 | 4 | 2 |
| Sugary Diet Intake<sup>a</sup> | < 3x/week | 3 | 4 | 1 |
|  | 3 – 6x/week | 4 | 1 | 4 |
|  | ≥ 1x/day | 3 | 5 | 5 |
| Last Dental Exam | < 1 year ago | 7 | 2 | 3 |
|  | 1-2 years ago | 1 | 1 | 1 |
|  | > 2 years ago | 2 | 7 | 6 |

<sup>a</sup> Subject level intake frequency was derived from a pre-defined list of dietary items commonly considered to be high in refined sugar. We assigned each participant to the highest frequency category reported across all items.

### Caries experience in SLB subjects

The DMFT and DMFS were significantly higher in both SLB groups relative to controls (p<0.05) (Fig. S1 & Fig. 1A). The severity of the carious lesions was also higher in SLB subjects, with most lesions being active, cavitated, and extending into dentin (i.e., ICDAS 5-6) (Table S3). Interestingly, tablet subjects had a higher DMFT and DMFS than film subjects, with more active carious surfaces extending into dentin or missing due to caries (p<0.05) (Fig. S1 and 1A). Strikingly, most of the carious teeth in the tablet group had 3 or more surfaces that were cavitated with active caries (Fig. 1B), meaning these teeth require extensive restorative treatment or even extraction.

**Figure 1:**
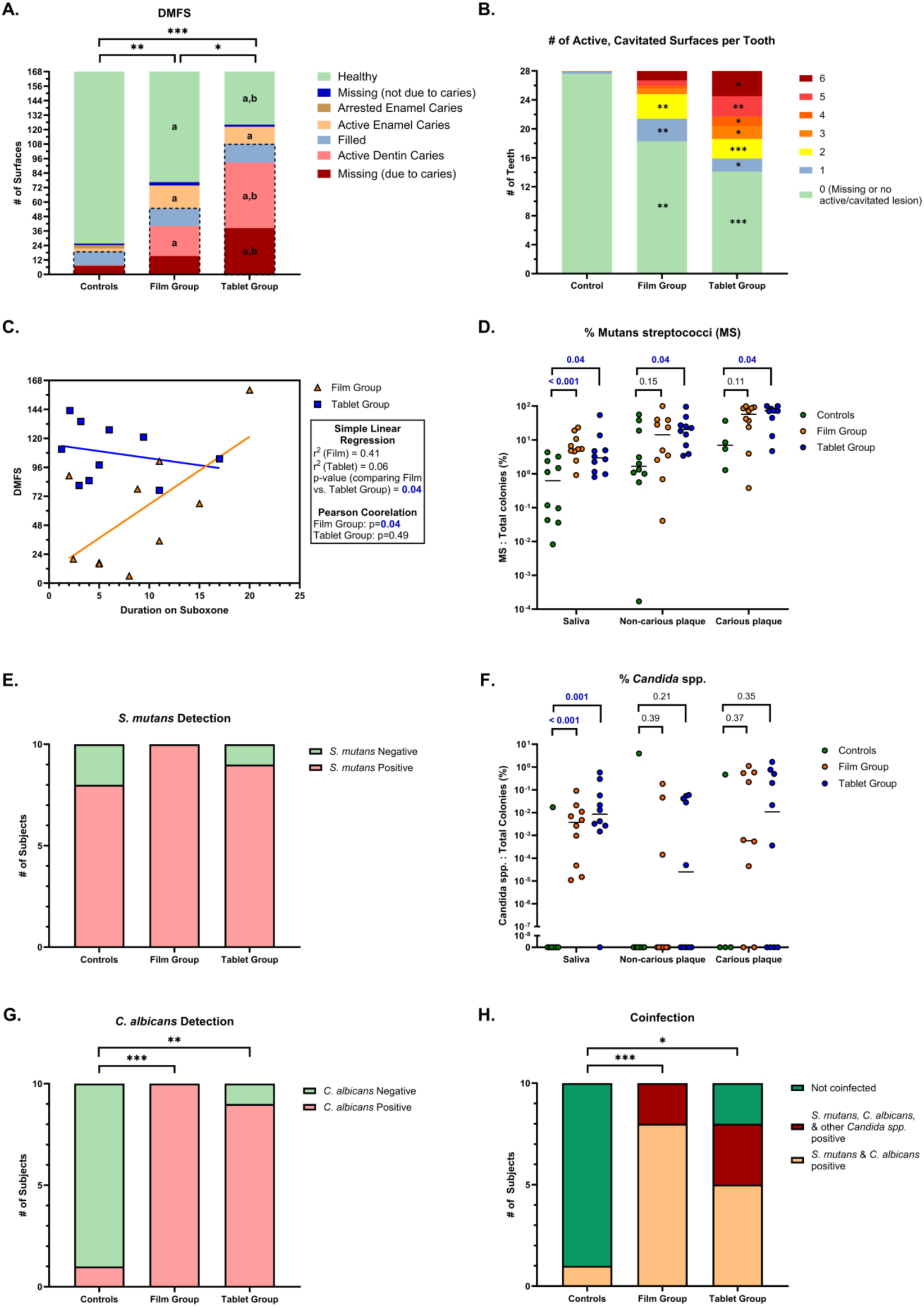
Caries experience and microbial burden of caries-associated microorganisms in patients taking SL buprenorphine. (A): Number of decayed, missing, or filled surfaces (DMFS) are outlined by black dashed lines. Sections are also separated by lesion severity (i.e., enamel vs dentin) and activity: (a) represents a statistically significant difference relative to control whereas (b) represents a statistically significant difference between the film and tablet group. (B) Number of teeth separated by number of active, cavitated carious lesions per tooth. (C): Correlation plot and analysis of DMFS vs duration taking SL buprenorphine films or tablets. (D): Relative percent burden of Mutans streptococci in each sample type. (E): Detection of *S. mutans* by subject using PCR. (F): Relative percent burden of *Candida* spp. by sample type. (G): Detection of *C. albicans* by subject using chromogenic agar. (H) Representation of the number of subjects co-infected with both *S. mutans* and *C. albicans*, or an additional *Candida* species. For numerical comparisons, FDR-adjusted p-values are reported and those bolded blue are statistically significant: *p < 0.05, **: p < 0.01, ***: p < 0.001.

Furthermore, we found that DMFS correlated with duration on SLB in film subjects (p=0.04), and a difference between the rates of DMFS in the film group and tablet group was observed (p=0.04) (Fig. 1C). This suggests the rate of caries onset or progression likely differs based on formulation.

### Burden of caries-associated microbes in SLB subjects

Subjects taking SLB tablets had higher levels of Mutans streptococci in saliva and dental plaque compared to controls (p=0.04), with the film group following a similar trend (Fig. 1D). Most subjects were *S. mutans* positive, with no significant differences between groups (Fig. 1E). Of note, only four subjects in the control group had a carious lesion with sufficient plaque to collect, limiting carious plaque sample size.

When compared to control, *Candida* spp. abundance was significantly elevated in saliva in both SLB groups (p≤0.001), but no significant differences were observed for dental plaque (Fig. 1F). Importantly, 90-100% of SLB subjects were positive for *C. albicans* compared to only 10% of the controls (Fig. 1G). When also accounting for those positive for *S. mutans*, the co-infection rate for SLB subjects ranged between 80-100% (Fig. 1H).

### Assessment of salivary risk factors in SLB subjects

There were no significant differences in unstimulated salivary flow rate (USFR) between groups (Fig. 2A). Interestingly, only one subject in the film and tablet groups had a USFR < 0.1 mL/min (diagnostic for clinical hyposalivation). When participants were asked about dry mouth frequency, half of the SLB subjects reported often or always (Fig. 2B). All SLB subjects were categorized as having a low or very low buffering capacity (Fig. 2C). Furthermore, baseline pH of both saliva and carious plaque in film and tablet subjects was lower than controls (p<0.01) (Fig. 2D). However, for non-carious plaque, only the film group displayed lower baseline pH (p=0.05) (Fig. 2D).

**Figure 2:**
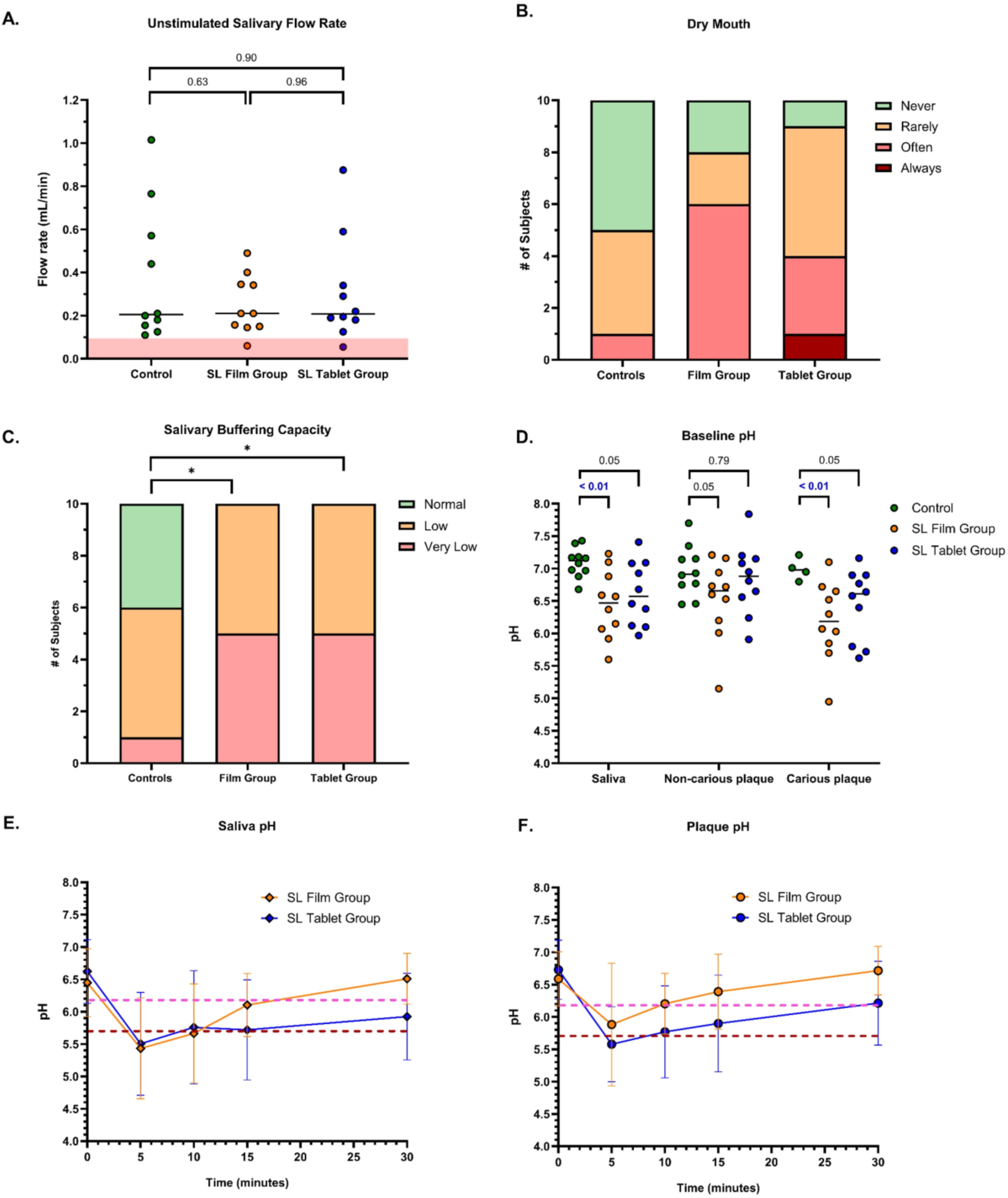
Critical salivary parameters and oral pH dynamics following SL administration of buprenorphine. (A) Each data point represents an individual subject’s unstimulated salivary flow rate, with the line representing the median. Multiple Mann-Whitney tests performed, and FDR adjusted p-values reported. The red area represents a flow rate < 0.1 mL/min which is representative of clinical hyposalivation. (B) Self-reported dry mouth and (C) salivary buffering capacity by category. Fischer’s exact test performed: *p < 0.05. (D) Baseline (i.e., resting) pH of all three sample types with each data point representing a single individual and the line representing the median. Multiple Mann-Whitney tests performed, and FDR adjusted p-values reported. Significant comparisons bolded in blue. (E-F) Oral pH kinetics (i.e., Stephan Curves) following SL administration. Each data point represents the mean ± SD. The dashed orange line represents the critical pH of dentin (pH 6.2) which the dashed red line represents the critical pH of enamel (pH 5.7).

Saliva and plaque pH kinetics following SLB administration were evaluated. Enamel and dentin demineralize at their critical pHs (5.7 and 6.2, respectively)^25^. A rapid decline below these values was observed, followed by a prolonged recovery phase (Fig. 2E-F), especially for the tablet group. The tablet group mean area under pH_5.7_ and time under pH_5.7_ from plaque was over 3-fold and 4-fold higher than the film group, respectively, although this difference was not significant (p=0.14) (Fig. S2A-B). The mean area and time under pH_6.2_ followed a similar trend (Fig. S2C-D), and the minimum plaque pH trended lower in tablet subjects relative to those using films (p=0.07) (Fig. S2E).

### Exposure of buprenorphine, its major metabolites, and naloxone in the oral cavity

Prior studies have proposed saliva acts as a reservoir for SLB^26,27^. We determined the salivary concentrations of buprenorphine, norbuprenorphine, norbuprenorphine-glucuronide, buprenorphine-glucuronide, and naloxone shortly after dosing (Fig. 3A-C). Mean concentrations of buprenorphine and naloxone remained above 10.0 μg/mL after 30 minutes (Fig. 3A-B), with no significant group differences. Norbuprenorphine concentrations tracked with the concentrations of buprenorphine, suggesting local phase I metabolism may be occurring (Fig. 3C). When observing concentration based on reported time since last dose, both buprenorphine and naloxone were drastically higher in saliva relative to their hypothetical plasma concentrations^28^ (Fig. 3D-E). Norbuprenorphine also remained elevated (Fig. 3F), however levels of norbuprenorphine-glucuronide and buprenorphine-glucuronide were undetectable at any point.

**Figure 3:**
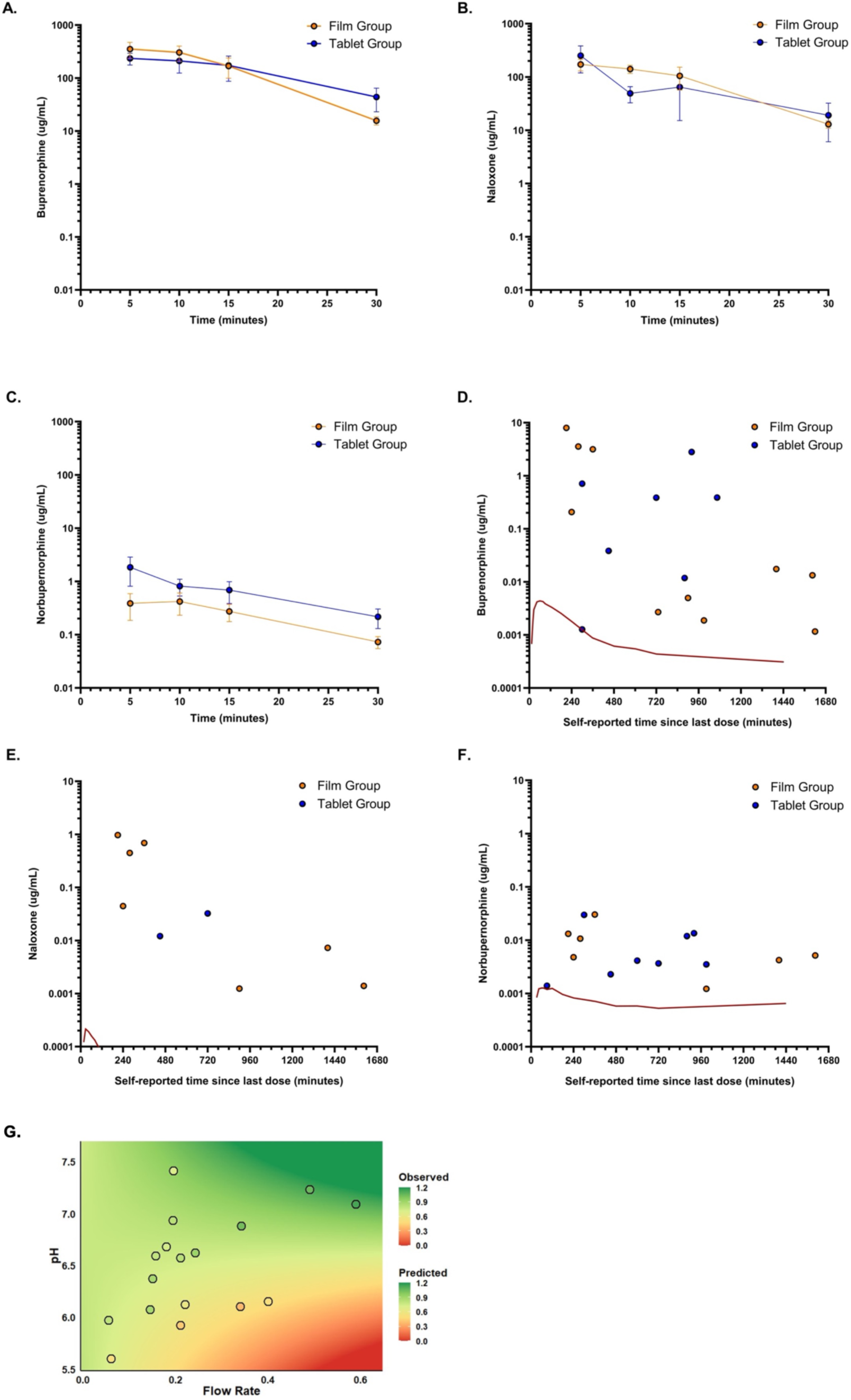
Salivary pharmacokinetics of buprenorphine, naloxone, and norbuprenorphine, and correlation with salivary parameters. (A-C): Each point represents the mean ± SEM salivary concentration after self-administration of sublingual dose. (D-F): Each point represents an individual participant. Subjects whose concentration was below the lower limit of quantification (LLOQ) are not shown. Red line represents plasma concentration-time profile following SL administration of Suboxone SL tablet (adopted from Dong et al., 2019). (G) Multiple linear regression analysis demonstrating relationship of the individual elimination exponent (*K_i_*) representing elimination of buprenorphine from saliva to resting pH and unstimulated salivary flow rate (USFR). Color of circle represents the observed *K_i_* value while background color describes the model-predicted value.

Because significant variability in buprenorphine salivary concentration was observed, we modeled individual concentration-time profiles using an empirical power-law model (Fig. S3). The individual elimination exponent, *K_i_,* representing the extent of elimination from saliva was determined. Multiple linear regression analysis revealed a significant correlation between pH and *K_i_* (p=0.003) (Fig. 3G). While no independent effect of USFR on *K_i_* was observed, a correlation was found when two-way interaction with pH was applied (p=0.02). Thus, our findings reveal that a lower pH increases buprenorphine retention in the oral cavity and is even more substantial in patients with lower USFR.

### Exploratory analysis of predictors of caries experience

We employed a random forest model to explore the major predictors of caries experience (DMFS/DS) in SLB subjects when considering known risk factors either collected by questionnaire or measured during the study. This model type has demonstrated accuracy in predicting caries risk^29^. Importantly, exposure to SLB emerged as a leading predictor of DMFS/DS, closely aligning with other well-established correlates of caries experience^30^ (Fig. 4A-B). While this cannot be interpreted as an independent predictor with causal inference, this finding is hypothesis-generating and further supports an association between SLB and dental caries. Furthermore, *C. albicans* and *S. mutans* abundance in saliva was also a strong predictor of caries experience, aligning with findings in other distinct patient populations^16,31^.

**Figure 4:**
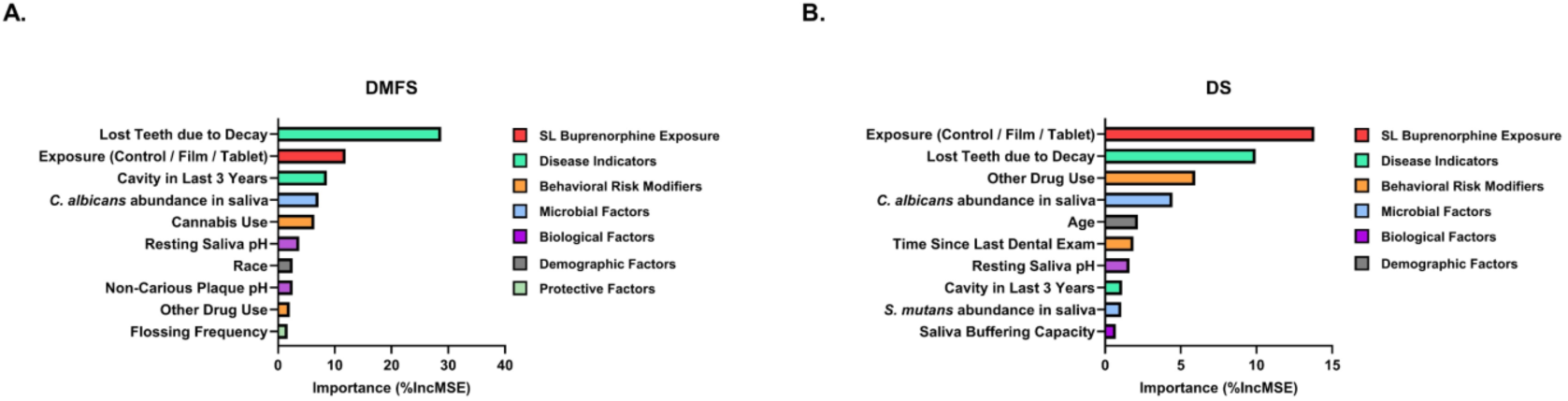
Major predictors of caries experience (DMFS/DS) in the study population. A unified random forest regression analysis was conducted to assess relative importance of each predicting variable measured by percentage increase in mean squared error (%IncMSE). The top ten predictors are represented above. Colors separate classifications of predictors.

## Discussion

Recent pharmacoepidemiologic and laboratory studies have demonstrated a clear association between SLB and dental caries, substantiating the FDA’s warning^5–12^. Nevertheless, studies assessing caries experience and determining the impact on several caries-associated microbiological and salivary risk factors were previously unestablished. This study revealed SLB subjects have elevated caries experience and elevated burden of the caries associated microbes *S. mutans* and *C. albicans*, with most SLB subjects being coinfected. SLB subjects have reduced resting pH and buffering capacity, with prolonged, elevated levels of buprenorphine and naloxone in saliva post-dose. By applying an empirical model and utilizing multivariate analysis, we found that the elimination exponent of buprenorphine correlated with the baseline salivary pH, suggesting an ion-trapping phenomenon contributes to oral retention of buprenorphine. Lastly, a random forest model demonstrated that exposure to SLB is a likely predictor of dental caries.

Interestingly, differences in caries experience between the tablet and film groups was observed. Factors that may explain this include differences in cumulative daily dose and acid exposure in dental plaque. While differences in these two parameters were not significant, they trended higher in tablet subjects. Recent *in vitro* studies have suggested that buprenorphine can upregulate *S. mutans* glucosyltransferase B in a concentration-dependent manner, thereby enhancing biofilm formation^12^. Therefore, increased doses may enhance *S. mutans* virulence and promote caries formation. Whether or not the presence of naloxone modifies this effect is a point for future investigation. Additionally, acid exposure trended higher in plaque from tablet subjects (p=0.14) and the minimum plaque pH trended lower (p=0.07). It is plausible that this formulation-dependent effect is due to differences in excipient composition^32^. While both tablets and films contain citric acid, the amount could vary. Furthermore, tablets also contain fermentable substrates such as lactose, mannitol, and corn starch, which can be converted to organic acids by cariogenic microorganisms localized within dental plaque, thereby inducing demineralization. The FDA defines a dietary item as cariogenic if it lowers plaque pH below 5.7 either during or within 30 minutes after consumption using wire telemetry^33^. While we did not use wire telemetry in this study, we did observe a pH drop below 5.7 in both film and tablet subjects, suggesting they meet these cariogenic criteria.

Intriguingly, we found a difference in the correlation of DMFS and duration on SLB between the film and tablet groups. The film group follows a positive trend (p=0.04) while the tablet group did not. Because we recruited subjects that have been taking SLB for 1+ years, it is plausible that an initial rapid increase in DMFS occurred in tablet subjects that was not captured in this study. Furthermore, rates of caries experience tend to plateau as DMFS is a cumulative indicator of caries burden^34^. All tablet subjects had an elevated caries experience, with >75% of teeth on average affected, and thus may also explain why the tablet group appeared to already reach a plateau and therefore lacks a strong correlation in caries experience over time. However, given the cross-sectional design, small sample size, and differences in treatment characteristics, this observation requires confirmation by a longitudinal study.

Prior research has demonstrated the importance of assessing *S. mutans* and *C. albicans* co-infection in severe caries^15,16,31^. Several mechanisms explain the intense synergism between these two oral microorganisms. *S. mutans* excretes lactic acid, which *C. albicans* can utilize as an energy source^35^. In turn, *C. albicans* protects *S. mutans* against environmental stressors, and they exhibit synergistic biofilm formation^36,37^. *Candida* prevalence typically exhibits higher positivity rates in children and the elderly with low rates in middle-age^38^. Intriguingly, most SLB subjects were middle-aged yet still exhibited extremely high rates of *S. mutans*-*C. albicans* co-infection. While it is unknown whether buprenorphine affects *C. albicans* pathophysiology, a prior study found that the opioid β-endorphin can induce germ tube formation *in vitro*, and may increase its virulence^39^. Opioids are also known to have an inhibitory effect on innate immune functions^40^, thus further investigations assessing the effects of opioids on salivary antimicrobial peptide production and oral mucosal immunity are warranted.

Saliva’s ability to neutralize acid prevents caries progression. Xerostomia has been proposed as a major factor in the association of SLB and dental caries^41^. A proposed mechanism is via opioid-induced release of gamma-aminobutyric acid (GABA), which can inhibit acetylcholine release at post-ganglionic parasympathetic fibers that innervate the salivary glands to stimulate saliva production^26^. However, clinical studies directly assessing salivary flow in SLB patients are lacking. Here, we found only two subjects taking SLB were clinically diagnosed with hyposalivation. While SLB does not appear to significantly impact flow rate, we found all SLB subjects had low or very low buffering capacity and lower baseline salivary pH, suggesting that buprenorphine, the excipients, or both may alter phosphate and bicarbonate buffering systems, thereby impairing neutralization and contributing to increased caries risk.

Previous studies have shown that buprenorphine exhibits prolonged exposure in the oral cavity^26,27^. A recent preclinical study demonstrated that SLB accumulates in the salivary glands where it can then undergo recirculation into the oral cavity^11^. Clearance from the oral cavity is primarily driven by swallowing rate, which correlates with salivary flow rate. Furthermore, buprenorphine is a weak base and therefore exhibits pH-dependent permeability, which may affect its mucosal distribution. Therefore, we employed multivariate analysis for correlation between pH, USFR, and buprenorphine elimination. The pH independently correlated with elimination, with a lower pH ion-trapping buprenorphine within the oral environment. While USFR did not correlate with elimination independently, it did have a significant effect as a two-way interaction with pH. This suggests that at lower USFR, the effect of pH on retention of buprenorphine is more substantial. Thus, a lower pH and USFR may compound caries risk by also prolonging elevated levels of buprenorphine within the oral cavity.

Following the FDA’s warning, some speculation attempted to attribute this association to a lack of proper oral hygiene, poor diet, and lack of dental care in OUD patients. However, pharmacoepidemiologic evidence suggests this speculation is indeed inaccurate. Etminan et al., demonstrated increased caries risk in OUD patients taking SLB versus naltrexone^7^. Barus et al., showed those taking SLB have increased caries risk versus those taking methadone^6^. We found that the mean DMFS for participants taking the SL film and tablet was 78.7 and 124.1, which is strikingly higher than previously reported for methamphetamine users (DMFS = 28.6) and heroin users (DMFS = 29.9)^42^. We also found frequency of oral hygiene practices did not differ between control and SLB groups. Exploratory random forest analysis revealed that exposure to SLB was a leading predictor of DMFS and DS for the entire study population. Taken together, our findings revealed this speculation as inaccurate and stigmatizing, further supporting the existing pharmacovigilance evidence suggesting a direct association between SLB use and increased caries risk.

## Limitations

Given a limited sample size, some comparisons may be underpowered and limit statistical inference. Accordingly, the results from the random forest analysis should be interpreted as exploratory, as small samples coupled with many correlated predictors can compromise model stability. However, this limitation does not negate the clinical relevance of the findings. As the first cross-sectional study assessing caries experience and associated microbiological and salivary risk factors in SLB patients, this study provides critical information to both addiction medicine and dental communities. These results can guide future longitudinal studies with larger cohorts to address confounders and enable subgroup analyses.

An additional limitation of this study is the lack of a disease-matched control group consisting of individuals with OUD who were not receiving SLB. Recruitment of this cohort was not feasible within our study population, as most individuals receiving treatment for OUD within the local community were prescribed SLB. Furthermore, those prescribed other medications approved to treat OUD (i.e., methadone or naltrexone) frequently received prior treatment with SLB, which would introduce additional potential confounding due to prior exposure. Additionally, studies suggest that methadone may independently increase caries risk^43^. Consequently, while the control group herein does not fully account for all OUD-related factors that may influence dental caries, it allowed for evaluation of caries experience and microbiological and salivary factors in patients without prior buprenorphine exposure and with similar demographics, oral hygiene practices, and intake of cariogenic dietary items.

Radiographs were not taken, so decayed teeth/surfaces may be underreported and caries experience underestimated. Furthermore, sites with erosion, attrition, or abrasion were not recorded. The examiner noticed several sites with erosion, particularly in the film group, indicating erosion should be documented in future investigations. Additionally, plaque pH was measured indirectly, potentially underestimating pH decline and acid exposure^44^. While wire telemetry has been shown to be more sensitive to pH fluctuations, it is costly, time consuming, and requires strict patient compliance. However, future studies should employ wire telemetry as it is likely to yield a steeper and prolonged pH drop, further exemplifying the cariogenic potential of SLB.

## Conclusions

Several factors may explain the association between SLB and dental caries. The marketed formulations themselves contain citric acid which reduces the local pH below the critical pH of enamel and dentin. These patients have a reduced resting pH and buffering capacity, thus lowering their ability to neutralize this acid. Chronic daily exposure favors highly aciduric microorganisms associated with dental caries, including Mutans streptococci and *Candida* species. Furthermore, acidic conditions increase retention of buprenorphine in the oral cavity, and elevated levels of buprenorphine may also promote biofilm formation by a keystone caries pathogen, *S. mutans*. Collectively, these risk factors create an ideal environment for rapid caries onset and progression.

## Data Availability

All data produced in the present study are available upon reasonable request to the authors

## Clinical Significance and Implications

SLB remains a highly effective, life-saving medication for OUD, and we do not advocate discontinuing treatment. Instead, caries-risk mitigation strategies should be integrated into care for patients receiving SLB. Close coordination between addiction medicine and dental providers may improve patient outcomes, and non-invasive preventive strategies including topical fluoride, xylitol-containing products, and oral hygiene instruction can substantially reduce caries burden in this high-risk population. Dental providers should also adapt their caries management approaches to best serve patients taking SLB.

## Key message

In this cross-sectional pilot study, sublingual buprenorphine was found to be associated with increased caries experience, likely driven by a combination of contributing microbiological and salivary risk factors. These findings support the current U.S. Food and Drug Administration (FDA) warning and underscore the need for future longitudinal studies with larger cohorts and additional comparator groups to assess for potential cause-effect relationships and provide additional mechanistic insight.

## Acknowledgements

We would like to thank all healthcare staff and providers at Meridian Behavioral Healthcare, Buprenorphine Treatment Centers, UF Health Florida Recovery Center, Gainesville After Hours Clinic, and New Season Treatment Center all located in Gainesville, Florida for their recruitment efforts.

## Ethical Approval

This cross-sectional clinical study was approved by the University of Florida Institutional Review Board (IRB202401848).

## Disclosure

The authors declare no conflicts of interest.

## Declaration of Generative AI Use

The authors report generative AI was not used in their research or preparation of this manuscript.

## Funding Sources

This work was supported by the University of Florida Research Foundation grant and the National Institute for Dental and Craniofacial Research Training grant (T90DE021990) and Fellowship grant (F30DE035751).

## Data Availability

All relevant data is presented within the main manuscript and supplemental materials. However, further details are available upon request.

## Notes

### Competing Interest Statement

The authors have declared no competing interest.

### Author Declarations

The University of Florida Institutional Review Board gave ethical approval for this work under the following protocol IRB20240184

