## supplemental data for "Sublingual Buprenorphine Is Associated with Increased Caries Experience and Altered Oral Microbial and Salivary Risk Factors"

### Supplemental Materials

#### Supplemental Methods

##### 1.0: Chemicals and Reagents

UPLC-MS grade formic acid, methanol, and acetonitrile (ACN) were purchased from Fisher Scientific (Waltham, MA, USA). The following certified reference standards were purchased from Millipore Sigma (St. Louis, MO, USA): phenacetin, buprenorphine, buprenorphine-glucuronide, norbuprenorphine, norbuprenorphine-glucuronide, buprenorphine-$d_4$, and naloxone.

##### 2.0: Bioanalytical Method Development and Sample Processing

##### 2.1 Instrumentation and chromatographic conditions

Chromatographic separation and mass spectrometric detection were conducted using an Acquity I-Class UPLC system coupled with a Xevo TQ-S micro triple quadrupole mass spectrometer (Waters Corporation, Milford, MA, USA). Separation was performed in Acquity UPLC BEH C18 column (100 mm × 2.1 mm, 1.7 $\mu$m), connected to a VanGuard BEH C18 pre-column (5 mm × 2.1 mm). The mobile phase consisted of (A) 0.1% formic acid in water and (B) 0.1% formic acid in ACN. The optimized gradient elution program was as follows: 0–0.8 min at 2% B; 0.8–2.0 min, gradient from 2% to 85% B; 2.0–3.0 min at 85% B; 3.0–3.1 min, gradient from 85% to 2% B; and 3.1–3.5 min at 2% B. The flow rate was set to 0.35 mL/min, with the column temperature maintained at 30 °C and the autosampler at 10 °C. The injection volume was 2 $\mu$L. The strong needle wash solution comprised water, acetonitrile, methanol, and isopropanol (1:1:1:1, *v*/v/*v*/v) with 0.1 % formic acid. The weak needle wash solution consisted of acetonitrile, methanol, and water (1:1:2, v/v/v) with 0.1 % formic acid. The seal wash consisted of water and methanol (95:5 v/v).

Triple quadrupole mass spectrometric analysis was conducted using electrospray ionization (ESI) in positive mode. For quantification purposes, multiple reaction monitoring (MRM)

For Peer Review

was utilized with selective mass transitions optimized for compound-specific parameters (Table S4). Nitrogen and argon served as the desolvation and collision gas, respectively. The desolvation temperature was held at 450.0 °C. This adjustment ensured that a minimum of 12 data points were collected across each chromatographic peak, providing sufficient temporal resolution for precise peak integration and quantification.

**2.2 Preparation of stock, standard, and quality control samples**

Stock solutions of buprenorphine, norbuprenorphine, buprenorphine-3-glucuronide, norbuprenorphine-3-glucuronide, and naloxone in were prepared in methanol to create the mother stock. Eight spiking stocks (10, 20, 50, 100, 200, 500, 1000, and 2000 ng/mL) were prepared by serial dilution in methanol from the combined mother stock. Calibration standards (CS) and quality control (QC) standards were prepared by combining 2 $\mu$L of each respective spiking stock solution with 18 $\mu$L of blank pooled human saliva. Eight CS were accurately prepared to cover a range of 1 - 200 ng/mL. Four QC samples were prepared at distinct concentration levels: 1 ng/mL as lower limit of quantification (LLOQ), 3 ng/mL as low-quality control (LQC), 90 ng/mL as medium-quality control (MQC), and 180 ng/mL as high-quality control (HQC). All stock and standard solutions were stored at − 20 °C until further use.

**2.3 Sample preparation and analysis**

Saliva samples from study subjects were prepared using protein precipitation method[1]. A quenching solution (QS) containing ACN with 0.1% formic acid was prepared. Two internal standards (IS), buprenorphine-d4 and phenacetin, were added to achieve a final concentration of 2 ng/mL in the QS. Buprenorphine-d4 was used as IS for buprenorphine and norbuprenorphine while phenacetin was used as IS for remaining analytes. A total of 180 uL of QS was added to 20 uL of freshly prepared CS and QC standards and clinical samples and were mixed thoroughly. Clinical samples were diluted in blank pooled human saliva when necessary to be within calibration range. Quenched samples underwent filtration using a Solvinert 96-well 0.45 $\mu$m filter

For Peer Review

plate by centrifuging at 2000 rpm for 5 min at 4 °C into a Waters 96-well sample collection plate (Milford, MA, USA). A cap mat was immediately placed, and samples were ready for injection.

Data acquisition was accomplished using MassLynx® software version 4.2, and TargetLynx® for data analysis. Calibration curves were constructed by plotting the peak area ratio of analytes to internal standard versus the nominal concentration of analytes. The calibration curve was fitted using least-square linear regression analysis.

**3.0: Application of empirical power-law model to saliva pharmacokinetics of buprenorphine**

In this study, we sought to determine if long-term retention of buprenorphine in the oral cavity was associated with either salivary pH or flow rate. Accordingly, the decline in salivary buprenorphine concentration was described using the following empirical power-law model:

$$C_{t,i} = C_{10,\,i} \cdot (\frac{t_{i,\,j}}{10})^{-K_i} \qquad (1)$$

$C_{t,i,}$ is the observed salivary buprenorphine concentration (µg/mL) for participant $i$ at time, $t_j$, (minutes);

$C_{10,i}$ is the individual predicted buprenorphine concentration (µg/mL) at 10 minutes;

$K_i$ is defined here as the individual exponent of elimination (unitless).

The 10-minute reference time was selected so that $C_{10,i}$ had a direct interpretation as the predicted concentration at an observed sampling time.

Taking the logarithm of both sides of equation (1) gives,

$$\log\left(C_{t,i}\right) = \log\left(C_{10,i}\right) - K_i \log\left(\frac{t_i}{10}\right) \qquad (2)$$



For Peer Review

Thus, $-K_i$ represents the slope of log concentration versus log time. A smaller $K_i$ indicates a less apparent decline in salivary concentration, which means increased retention in the oral cavity. Therefore, the exponent of retention, $K_i$, is not a rate constant but rather describes the rate at which concentration declines when drug washout is governed not by a single first-order elimination process, but by a broad distribution of residence times across heterogeneous microenvironments. Because $K_i$ is dimensionless, it characterizes the structural and diffusional complexity of elimination from the oral cavity rather than a classic kinetic rate constant typically described in pharmacokinetics.

A multivariate analysis assessing the correlation between resting pH, unstimulated salivary flow rate, and $K_i$ was performed as described in the main manuscript.

For Peer Review

#### Supplemental Tables and Figures

**Table S1: Inclusion and exclusion eligibility criteria for the study population.**

| Inclusion Criteria | Exclusion Criteria |
|---|---|
| 18-90 years of age | Use of any antibiotics, antifungals, or steroid treatments (including inhalers) within the past month |
| Must have at least 10 natural teeth remaining | Pregnant or breastfeeding |
| Diagnosis of opioid use disorder **(Omit for healthy controls)** | Autoimmune conditions or any severe systemic conditions (ASA IV or more) |
| | Medical conditions requiring any type of pre-medication or medication modification prior to dental treatment (ie. bleeding disorders, artificial heart protheses, joint replacements) |
| Taking sublingual tablets or films containing buprenorphine for at least 1 year **(Omit for healthy controls)** | Use of a removable prothesis (ie. partial dentures, complete dentures, orthodontic appliances, occlusal guard, whitening trays, Essix retainers) |
| | Use of a toothpaste or mouthwash containing either prescription strength fluoride or hydrogen peroxide within the past month (standard over-the-counter toothpastes and mouthwashes are acceptable) |
| | Use of prescribed chlorhexidine mouth rinse or any other prescribed antimicrobial mouthwash within the past month |
| | Prior radiation to the head or neck |

For Peer Review

**Table S2: Summary of results for buprenorphine use-related questions**. A total of 31 subjects completed the questionnaire. One film subject was excluded from analysis due to recently switching from film to tablet, except for disintegration time a different film subject was excluded as they prematurely swallowed the film following self-administration. Therefore, a total of 10 subjects per group were analyzed. Mean (SD) are reported for numerical variables and number of subjects are reported for categorical variables. Mann-Whitney test performed for numerical variables and Pearson chi-square test performed for categorical variables: *: p-value < 0.05; **: p-value < 0.01.

| | | Film Group | Tablet Group |
|---|---|---|---|
| Cumulative daily dose, mean (SD), mg | | 18.1 (8.0) | 23.2 (4.5) |
| Co-administered with Naloxone** | Yes | 10 | 3 |
| | No | 0 | 7 |
| Drug Product Preference* | None (only taken one formulation) | 1 | 3 |
| | No preference | 0 | 1 |
| | Prefers Films | 9 | 2 |
| | Prefers Tablets | 0 | 4 |
| Duration since initiating SL buprenorphine treatment, mean (SD), years | | 8.83 (5.68) | 6.19 (4.91) |
| Self-reported change in dental condition since initiating buprenorphine treatment | Improved | 0 | 0 |
| | Not changed | 2 | 1 |
| | Slightly worsened | 2 | 3 |
| | Drastically declined | 6 | 6 |
| Disintegration time, mean (SD), minutes | | 9.9 (6.0) | 10.8 (7.6) |
| Taste Rating | Excellent | 0 | 0 |
| | Acceptable | 2 | 3 |
| | Poor | 3 | 3 |
| | Very Poor | 5 | 4 |
| Awareness of FDA Warning | Yes | 4 | 6 |
| | No | 6 | 4 |

For Peer Review

**Figure S1: Number of decayed, missing, or filled teeth (DMFT).** Dashed black lines represent categories included in the overall DMFT calculations. Multiple Mann-Whitney Tests performed; (a) represents a statistically significant difference relative to controls whereas (b) represents a statistically significant difference between the film and tablet groups. FDR-adjusted p-values were calculated; *: $p < 0.05$, **: $p < 0.01$, ***: $p < 0.001$.

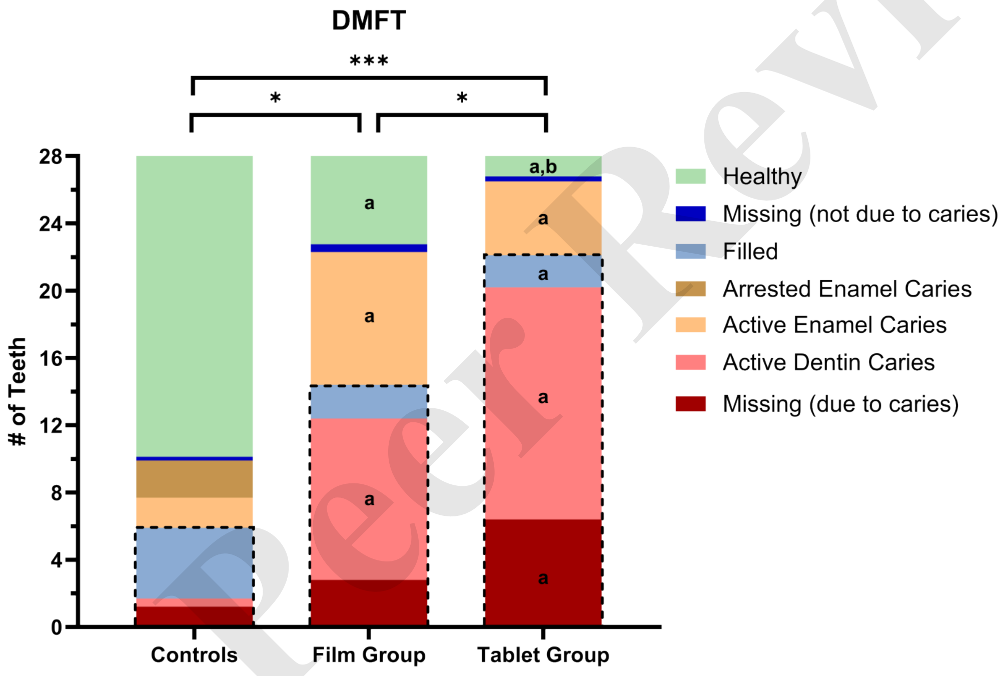


For Peer Review

**Table S3: Number of carious teeth and surfaces by severity and activity.** Mean ± SD listed per group. Severity categorized based on ICDAS-II criteria. No significant differences between film and tablet groups found.

| | Arrested Carious Teeth | | | Active Carious Teeth | | |
|---|---|---|---|---|---|---|
| ICDAS Score | Controls | Film Group | Tablet Group | Controls | Film Group | Tablet Group |
| 1-2 | 2.20 ± 3.49 | 0 ± 0[a] | 0 ± 0[a] | 1.80 ± 2.15 | 7.90 ± 5.09[a] | 4.40 ± 3.27 |
| 3-4 | 0 ± 0 | 0 ± 0 | 0 ± 0 | 0.40 ± 0.97 | 0.40 ± 0.52 | 0.30 ± 0.48 |
| 5-6 | 0 ± 0 | 0 ± 0 | 0 ± 0 | 0.10 ± 0.32 | 9.20 ± 7.15[a] | 13.50 ± 5.19[a] |
| 1-6 | 2.20 ± 3.49 | 0 ± 0[a] | 0 ± 0[a] | 2.30 ± 2.71 | 17.50 ± 6.74[a] | 18.20 ± 4.32[a] |
| | **Arrested Carious Surfaces** | | | **Active Carious Surfaces** | | |
| ICDAS Score | Controls | Film Group | Tablet Group | Controls | Film Group | Tablet Group |
| 1-2 | 2.80 ± 4.71 | 0 ± 0[a] | 0.10 ± 0.32 | 2.40 ± 3.03 | 17.50 ± 9.16[a] | 14.20 ± 9.59[a] |
| 3-4 | 0 ± 0 | 0 ± 0 | 0 ± 0 | 0.40 ± 0.97 | 2.40 ± 1.71[a] | 2.50 ± 2.51[a] |
| 5-6 | 0 ± 0 | 0 ± 0 | 0 ± 0 | 0.10 ± 0.32 | 23.80 ± 27.31[a] | 51.70 ± 28.54[a] |
| 1-6 | 2.80 ± 4.71 | 0 ± 0[a] | 0.10 ± 0.32[a] | 2.90 ± 3.70 | 43.70 ± 31.95[a] | 68.40 ± 26.01[a] |

[a] Statistically significant difference relative to control group.

For Peer Review

**Figure S2: Assessment of caries associated pH parameters by group**. All parameters were determined using GatorpH software by group and sample type (i.e., plaque or saliva). Violin plots represent the distribution, with the solid black line within each plot representing the median. Mann-Whitney tests performed to compare groups within sample type; FDR-adjusted p-values reported. (E) Pink dashed line represents critical pH of dentin (pH 6.2) and red dashed line represents the critical pH of enamel (pH 5.7).

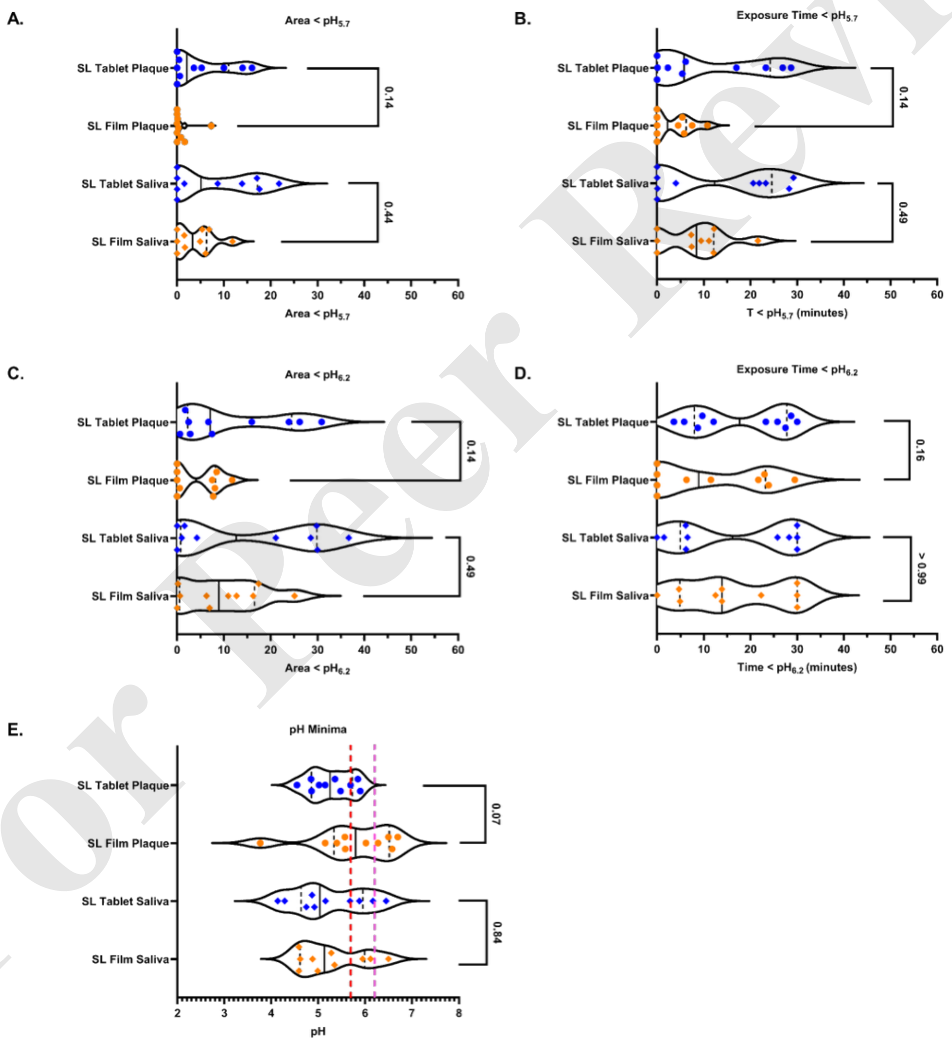


For Peer Review

**Table S4: Summary of mass spectrometry compound parameters**. IS: Internal standard.

| Analyte | Mass Transition | Capillary Voltage (kV) | Cone Voltage (V) | Collision Energy (V) |
|---|---|---|---|---|
| Buprenorphine | 468.3 → 55.1 (Quantifier) | 0.50 | 34 | 54 |
| | 468.3 → 101.0 (Qualifier) | 0.50 | 34 | 44 |
| Norbuprenorphine | 414.1 → 83.06 (Quantifier) | 0.50 | 72 | 46 |
| | 414.0 → 101.0 (Qualifier) | 0.50 | 72 | 32 |
| Buprenorphine-3-glucuronide | 644.3 → 468.4 (Quantifier) | 0.50 | 2 | 38 |
| | 644.3 → 55.1 (Qualifier) | 0.50 | 2 | 80 |
| Norbuprenorphine-3-glucuronide | 590.3 → 414.3 (Quantifier) | 0.50 | 30 | 40 |
| | 590.3 → 101.1 (Qualifier) | 0.50 | 2 | 58 |
| Naloxone | 328.1 → 212.1 (Quantifier) | 0.50 | 20 | 42 |
| | 328.2 → 253.2 (Qualifier) | 0.50 | 4 | 22 |
| Buprenorphine-d4 (IS) | 472.3 → 59.0 | 0.50 | 6 | 58 |
| Phenacetin (IS) | 180.1 → 110.0 | 0.50 | 34 | 20 |

For Peer Review

**Figure S3: Individual fit of buprenorphine concentration-time profiles in saliva.** Each plot represents the observed concentration-time profile for each subject. Subject F2 was excluded due to swallowing the film prior to complete disintegration. Each point is the observed concentration at 5, 10, 15, and 30 minutes. Any point past 30 minutes represents the concentration of the pre-dose sample extrapolated based on self-reported time since last dose. Each line represents the individual power-law model fit. All subjects self-administered an 8 mg dose of buprenorphine sublingually except subject F5 received a 4 mg dose and subjects F4 and F9 received a 2 mg dose. The plots were generated in Monolix Suite 2024 R1 (Lixoft, Antony, France) and in R (version 4.6.0) with RStudio (version 2026.04.0).

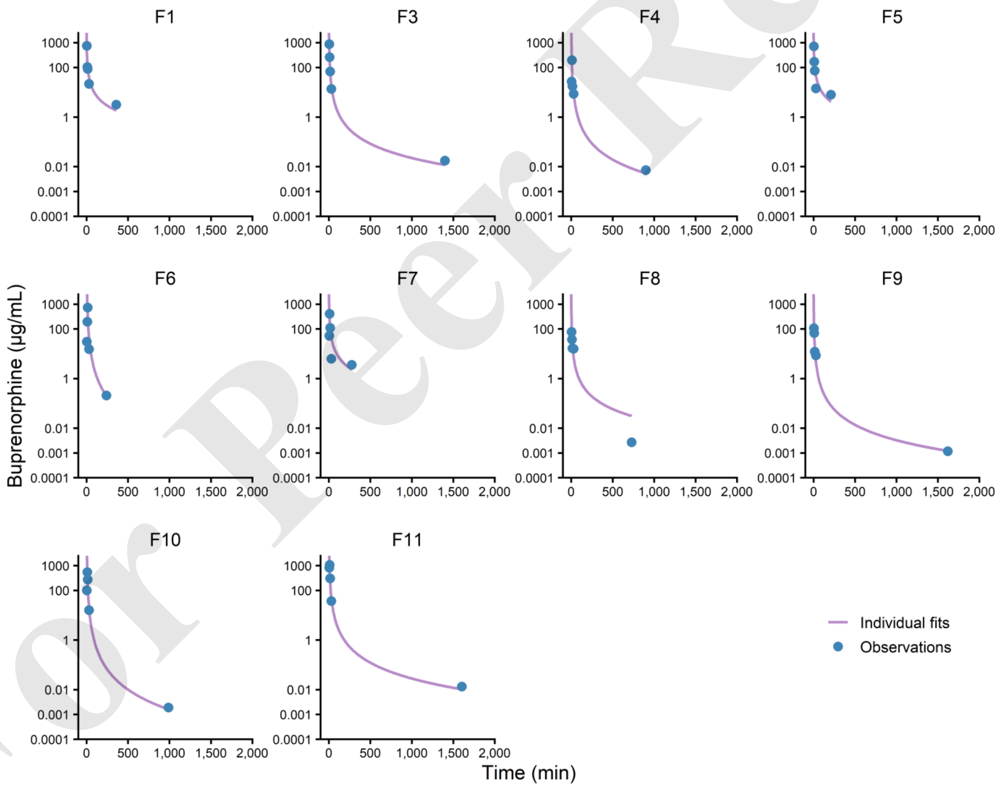


For Peer Review

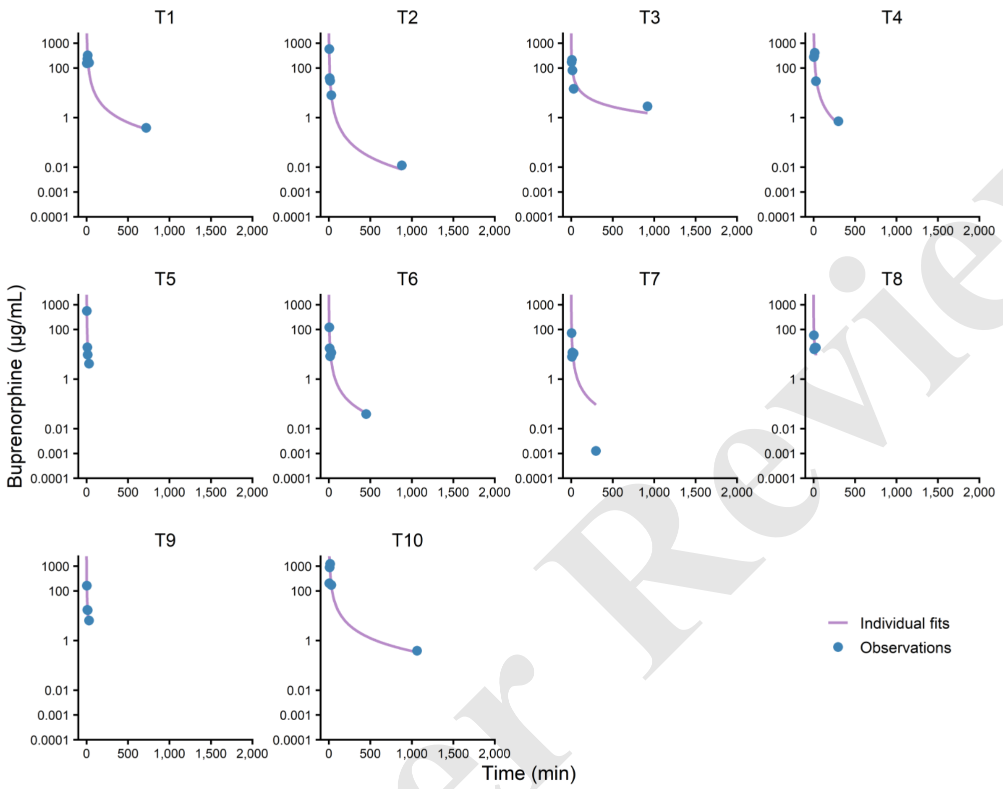


For Peer Review

For Peer Review
